# Healthcare workers’ beliefs, practices and experiences regarding the COVID-19 pandemic and resulting governance from preparedness and response plans in Faranah, Guinea

**DOI:** 10.1101/2025.06.10.25329321

**Authors:** Carlos Rocha, Moussa Douno, Lena Landsmann, Kamis Diallo, Rebekah Wood, Mardjan Arvand, Carolin Meinus, Mamadou Diallo, Almudena Marí-Sáez

## Abstract

There is a growing interest in the implementation of people-centered components within outbreak preparedness and response plans and their resulting governance. Information on this topic still remains scarce, in particular in settings like Guinea, West Africa. We draw from 21 qualitative interviews with healthcare workers in the context of COVID-19 from the Faranah Regional Hospital in Guinea to explore pandemic preparedness and response adherence as well as its corresponding senses of vulnerability and risk, information sources, perceived information reliability, associated rumors and previous outbreak experiences. To achieve a broader analysis, we make use of the “therapeutic landscape” concept which conceives health provision as a process constructed through practices, beliefs and experiences. A theme articulated across interviews was an overabundance of information, which caused confusion mainly due to rumors and a lack of trust in the country’s health system. Initial fear gradually diminished due to the disease’s low mortality rate in the country. HCWs described preparedness enforcement in the city of Faranah that started strong but lightened overtime. This waning of enforcement marked a difference between the city and the hospital. HCWs appraised the first positive case declaration in the city as the most significant moment, followed by a strong community reaction hindering the pandemic governance. We conclude that preparedness and response plans need to better socially situate their interventions and devote more structural efforts into incorporating the social landscapes of a disease and an outbreak. Incorporation of these social landscapes facilitates understanding of operational barriers to their people-centered approaches. Responses should therefore comprehend 1) hospitals as applying different pandemic understandings transcending biomedical and scientific orders and 2) HCWs as portraying shifting pre-existing identities leading to marked in-/out-group distinctions directly influencing healthcare, risk perception as well as information and rumors management.

## Introduction

In the aftermath of the 2014 – 2016 Ebola Virus Disease (EVD) outbreak in West Africa, there has taken place a growing involvement of the social sciences in outbreak preparedness and response not only in the African continent but worldwide (1–4). The COVID-19 pandemic has further highlighted the importance of a context-integrated approach to prevention, preparedness and response (5). Context-specific and community centered approaches are considered key to comprehend local understandings of diseases, identify multisectoral drivers of vulnerability, strengthen local resilience networks and develop effective models for engaging communities in and beyond the context of COVID-19 (6–8).

International and national preparedness and response plans for the African continent emphasized the need to engage communities around COVID-19 by reinforcing capacity and local solutions from community leaders, civil society groups and healthcare workers (HCWs) (9). Despite this increasing awareness, information still remains scarce on the actual implementation and governance consequences of context-specific and community centered approaches (10, 11).

In Sub-Saharan Africa, while disastrous effects related to COVID-19 were initially expected due to a fragile healthcare system structure, countries recorded lower infection and mortality rates than most countries in Europe, America and Asia (12–14). Nonetheless, the impact on health and economic systems, as well as livelihoods and citizen-state relationships, has been considerably high (15). This fact highlights the need to carry out research able to provide depth into these structural tensions, and how they feedback into national and international preparedness and response plans (16, 17).

At the beginning of the pandemic in Guinea, marked by its first case in March 2020 (18), the National Preparedness Plan included the dissemination of appropriate information on the disease, its mode of transmission and prevention. The National Response Plan sought to increase the sytems’s capacity and communication through awareness raising, radio and television broadcasts, and the production of communication tools as well as community engagement (19). The response strategies recruited more HCWs, but Infection Prevention and Control (IPC) training on COVID-19 was lacking or unsatisfactory (20). Among the country’s HCWs, rapid situational assessments showed a marked epidemiological knowledge disparity between the capital (Conakry) and the interior regions, where knowledge levels were lower (21, 22). Social media and traditional media (radio and television) were identified as the primary information source for HCWs both in rural as in urban settings (21, 22).

In a post-Ebola context, with the population demonstrating strong suspicion of the health system (23–25), the COVID-19 pandemic may have propagated already problematic pre-existing perceptions of HCWs (e.g. perceived as sources of contagion) (26, 27). The occurrence of this new pandemic may have also contributed to a maintained distrust and further decrease in the use of health services, exacerbating inequitable patient care and hospitalization conditions (28). Nonetheless, the country - in particular its peri-urban and rural areas - had relatively lower reporting of positive COVID-19 cases, morbidity and mortality as compared to global rates (28, 29).

HCWs are at the frontline of the response to infectious diseases outbreaks. Low perceived preparedness for COVID-19 response has been associated with stress and burnout due to mental stress, physical fatigue and separation from families and loved ones (30–32). Structural challenges in the health system related to inadequate infrastructure, lack of universal coverage and poor working conditions make the HCWs’ work environment deficient under unhealthy and corrupt therapeutic relationships (33–36). These aspects place healthcare facilities at risk of becoming outbreak or mistrust hotspots in pandemic conditions(37).

More research is needed to provide evidence on local implementations of response and preparedness plans. A comprehensive understanding of health facilities is necessary to situate hospitals and the practices within as products of social, political, and biomedical factors which shape disease transmission (38–40). This in-depth understanding allows to complexify HCWs’ experiences and the role expected from them by preparedness and response plans (41). To substantiate the HCWs’ role within the response to the COVID-19 pandemic, we make use of the “therapeutic landscape” concept in order to approach health provision as a process that is lived, understood and constructed through actions of a group of people in a particular place (42–44).

This study aims to explore some of the emergent features of the Faranah Regional Hospital’s therapeutic landscape by analyzing the beliefs, practices and experiences of its HCWs vis-à-vis the preparedness and the response for COVID-19 at the hospital and in the city of Faranah.

## Materials and methods

### Ethics approval

Ethical approval (approval number: 036/CNERS/21) for the study was granted by the National Health Research Ethics Committee of Guinea (*Comité National d’Éthique pour la Recherche en Santé)*.

### Study design and study setting

We conducted a three-month qualitative study from the 1^st^ of August to the 31^st^ of October, 2021, composed of semi-structured interviews (IDIs) with HCWs of the Faranah Regional Hospital.

The Faranah Regional Hospital is a public hospital in the Upper Guinea Region. It employs 97 HCWs and serves as a referral hospital for the Faranah region, covering a population of more than 300.000 which belongs mainly to the *Malinké* ethnic group. The hospital consists of 16 wards, including a surgery, laboratory, maternity ward and a Center for the Treatment of Epidemics (CT-EpI).

### Participant selection

All HCWs were eligible to participate in the study. We used convenience sampling aiming at a representative distribution of age and professional groups and an equal distribution between genders. Following a snowball methodology, each participant was asked to recommend a colleague who could be interested in participating in the study. Participants under the age of 18 years and those unwilling or unable to consent were excluded. Written informed consent in French was obtained from each participant.

### Data collection

With the HCWs who consented to participate, a meeting was scheduled for a day, time and place that made them most comfortable. Data was collected at the Faranah Regional Hospital. Each interview session lasted between 22 and 75 minutes. The interviewer was already known to the participants from previous research carried out at the hospital (45). Interviews were held mainly in French and/or other local languages familiar to the interviewer. The interviewer translated the data if needed, transcribed verbatim and analyzed in French. To follow the correct development of the interview, the interviewer took fieldnotes that served to ensure question quality, appropriateness and data saturation, which was achieved when no more new information was coming up in each of the interview guide topics. Quotations were translated into English for the purposes of this paper.

### Interview guide

We developed a semi-structured interview guide which included questions about the pandemic progression over time, the sources and changes in the type of information about it as well as associated memories and emotional appraisals. The guide also covered opinions and mental images about the virus and the pandemic as well as questions about the perception of the changes caused by the enforcement of preparedness and response strategies in the hospital, the Faranah Region and the country.

### Data analysis

We performed an inductive coding on a random selection of transcripts. This resulted in the creation of a codebook which was then applied to all of the data using NVivo 12 qualitative data analysis software. Following a grounded theory approach, we carried out a subsequent inductive analysis through an iterative process allowing the inclusion of new theoretical frameworks. The theoretical framework of hospitals as therapeutic landscapes was identified as the most appropriate as it has been applied in similar contexts to analyze health care provision (43). It views hospitals as both material and symbolic structures creating and created by social forces, some anchored in historical processes like the bureaucratic organization of healthcare provision from the colonial to the post-independence administration (46, 47). Furthermore, the concept acknowledges that healthcare provision acquires social meaning through the habits and beliefs of the people that practice it (44). The HCWs’ identity incorporates the clinical/medical practice (e.g. profession or years of experience) and community social leadership as well as worldviews (e.g. magical or religious) that are intrinsic to health care provision (37).

## Results

We interviewed 21 HCWs from different hospital services (general medicine, surgery, radiology, maternity, CT-EpI, among others) and professions. We outline participant characteristics in Table I.

**Table I.**
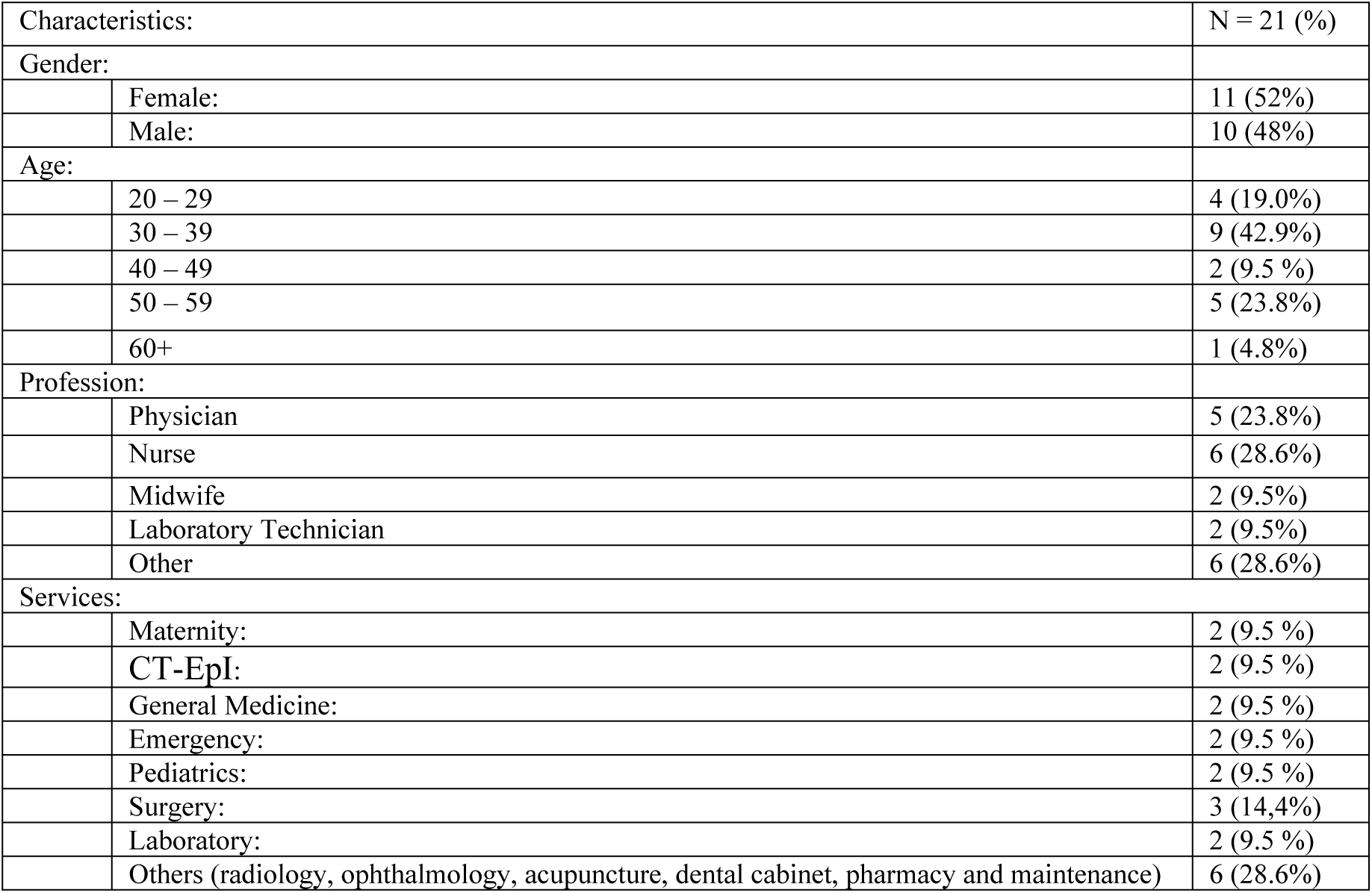
Participants’ characteristics (N=21)

| Characteristics: |  | N = 21 (%) |
| --- | --- | --- |
| Gender: |  |  |
|  | Female: | 11 (52%) |
|  | Male: | 10 (48%) |
| Age: |  |  |
|  | 20 – 29 | 4 (19.0%) |
|  | 30 – 39 | 9 (42.9%) |
|  | 40 – 49 | 2 (9.5 %) |
|  | 50 – 59 | 5 (23.8%) |
|  | 60+ | 1 (4.8%) |
| Profession: |  |  |
|  | Physician | 5 (23.8%) |
|  | Nurse | 6 (28.6%) |
|  | Midwife | 2 (9.5%) |
|  | Laboratory Technician | 2 (9.5%) |
|  | Other | 6 (28.6%) |
| Services: |  |  |
|  | Maternity: | 2 (9.5 %) |
|  | CT-EpI: | 2 (9.5 %) |
|  | General Medicine: | 2 (9.5 %) |
|  | Emergency: | 2 (9.5 %) |
|  | Pediatrics: | 2 (9.5 %) |
|  | Surgery: | 3 (14,4%) |
|  | Laboratory: | 2 (9.5 %) |
|  | Others (radiology, ophthalmology, acupuncture, dental cabinet, pharmacy and maintenance) | 6 (28.6%) |

We present the results under the following three categories dedicated to beliefs, practices, and experiences.

### HCWs’ beliefs about COVID-19

Defining a belief as a precept accepted as true or real (48), this section presents types and reliability of information sources, as well as perceptions about rumors and risk.

#### Information content and sources

Participants explained receiving information about the appearance of a new and highly contagious virus responsible for a severe respiratory syndrome originating in China. The information included topics like the nature of the virus, virulence, its prevention and control. One theme that HCWs consistently articulated was the case fatality rate, which they qualified as high and thus unsettling. The primary sources of information were the media. Overall, participants mentioned social media (mainly Facebook), television, radio and the Google search engine. In addition, a few participants explained getting further information at the Faranah Regional Hospital through the director and department heads during staff meetings.

#### Reliability of information sources

While some of the interviewed HCWs qualify radio and television as reliable to learn about the pandemic, others found that the most credible information sources are the websites from e.g. the WHO, UNICEF or OIM, the communiqués from the Ministry of Health or the National Agency for Sanitary Security (ANSS) and information from the hospital directives. One participant summarized this opinion by stressing the reliability of leadership figures: ‘the good information is obtained through […] doctors, it is the doctor who must give the right information so that we can better avoid this disease and raise awareness to improve the health of the population’ (IDI 19).

Some argued that the most trustworthy source of information was the Guinean state television that broadcasts ANSS “communiqués” every evening. However, the latter was questioned by others since the community death counting was perceived as flawed. Because of this, one HCW reported that ‘the international channels are especially reliable, because […] they tell the exact figure per day, the number of deaths, the number admitted in resuscitation (ICU)’ (IDI 14). For these reasons, the international media elicited credibility, considering that ‘the statistics they put out are not biased’ (IDI 14) since they report the exact numbers of cases and deaths.

#### Conflicting and reassuring information

Contradictions about tests results were especially prominent when enquired about conflicting information. As one HCW said: ‘sometimes we tell them today [to the patients], you’re positive, after a few hours, we call them and tell them, you’re negative, so that’s a contradiction’ (IDI 4). HCWs argued that this is a source of considerable confusion both for them and patients since it leads to questioning the reliability of antigen rapid tests, which they describe as being sensitive to many other viruses.

Another participant explained that many patients underestimate the danger of the disease by equating it with influenza or malaria. In this sense, they claimed that it already existed in the country or that the Guinean population has antibodies. One HCW explained: ‘for example in Conakry, it is said that the disease is not true, others even say that it is “corona-palu” [alluding to the word ‘paludism’, another denomination for Malaria in francophone West Africa]’ (IDI 6).

All participants stressed that there was also reassuring information. For the interviewed HCWs the most comforting was the high recovery rate among admitted patients in Guinea. They also reported that low case fatality rates gave them hope that a treatment would eventually exist. In addition, they found the information on preventive measures as reassuring because ‘when they are respected in the services and the families, there will be no contamination. What is also reassuring for me is when, out of 100 sick people, we say that 80 to 90 have been cured, or out of 100 people, there have only been 2 deaths, that is really reassuring’ (IDI 9). They also perceived information on the development of vaccines as encouraging.

Other participants felt reassured with the pandemic’s progression in Europe and North America,

> “because Africa is a developing continent, some people think that [epidemics] are only for Africans, because we have nothing. When I find that the same problems exist in highly developed countries, people are dying, it allows me to conclude that it is a global problem” (IDI 13).

#### Risk perceptions

The risk perceptions presented by the HCWs were sometimes in contradiction with the information gained from the biomedical practice (detailed above). One theme that participants consistently articulated is the perception of high risk brought by this disease. The worldwide high mortality rate, the contagiousness and the economic and socio-cultural consequences are held accountable for this elevated risk perception. They also associated this with the fact that the pandemic is a significant problem worldwide. As one HCW put it:

> ‘from the moment I saw [information on COVID-19 on] these continents [i.e. Europe and North America], I kept seeing the number of corpses, the number of deaths, and I was afraid, I said, listen, if the people we rely on are in trouble like this, if it happens in Africa, what will happen?’ (IDI 10).

Fear and sadness were the most prominent emotions evoked about the received information. These emotions were elicited by the awareness that this disease had foreign origins and was being brought to the country by people able to travel. One HCW explained: ‘I was really scared. Especially of the people who have traveled outside of the country. Even my son is abroad, he even wanted to come […] but as soon as the problem started, I talked with him and I told him not to come, to stay there’ (IDI 3). Furthermore, as one participant put it, ‘once you are told that the ministers are attacked, the senior staff is attacked, you must be afraid since the disease has not spared the authorities’ (IDI 12).

Participants described a change in the perception as the COVID-19 pandemic evolved. Several described the initial fear to gradually diminish mainly due to the low mortality rate in Guinea, the decrease in the number of positive cases and the high recovery rate among hospitalized patients.

#### Rumors about COVID-19

Overall, participants reported hearing community members denying the presence of the virus in Guinea, and even the actual emergence of a new and deadly disease. As such, participants noted that there had been various rumors and conspiracy narratives about this new infection. According to them, rumor sources include “word of mouth”, the radio, print and social media. Participants consistently linked the rise of these rumors and conspiracy narratives with the country’s political situation as the pandemic coincided with 2020 presidential elections. The president in office sought, with success, to run for a third term.

Almost all participants perceived these rumors and conspiracy narratives as untruthful. Yet, they do not report any effort undertaken by the authorities or the Faranah Regional Hospital to dismantle these rumors. HCWs explained that the elevated illiteracy level of the Guinean population facilitated their rapid spread. One participant argued that ‘the high amount of contradictory information is quite normal, living in an environment where the illiteracy rate is so high. 80% of the population says that the disease doesn’t exist, that it’s invented’ (IDI 10). Other participants emphasized social and religious factors such as inter-ethnic tensions. One HCW understood the emergence of this disease as a punishment from God because of humanity’s trespasses: “Such a disease came. Is it not a sanction of God? […] in Italy they allowed in a church the homosexual marriage. God doesn’t like that” (IDI 7).

### Practices related to COVID-19

We define a practice as the application of an idea or a belief by the actions of social actors within given societies (48). This section presents perceptions about the enforcement of response plans and their implementation as well as strategies to counteract vulnerability and rumors.

#### Counteracting vulnerability and rumors

All the participants reported that the means for feeling protected against this pandemic are the infection prevention and control measures; especially hand hygiene, mask wearing and compliance with social distancing. HCWs estimated community engagement as effective to counteract rumors and enforce IPC.

All HCWs reported having heard of non-biomedical treatments for this new infection as well as the use of medications designed to treat other infectious diseases. Mostly, the treatments they heard about were herbal, including artemisia, cinchona bark, lemon, garlic, acacia, ginger, and other local plants they named in the local language: “Popa” and “Sindjan lilin”. Some participants also mentioned that the idea that the virus is susceptible to bitter plants was widespread, motivating people to drink the cinchona-based decoctions and/or “Popa”. While some participants testified having seen acquaintances using some of these plants, only one HCW claimed to have bought, prepared and drunk one with their children at home. This participant acknowledged: ‘I advised people, even my children who are in Conakry, to boil and drink it’ (IDI 7).

Most of the HCWs stated they did not believe in the decoctions and other non-biomedical treatments. One HCW explained: ‘these treatments I can say no, personally, as a healthcare worker, I do not prescribe them’ (IDI 4). However, some were reluctant to admit whether they believed in these non-medical treatments or not, stating that what is said should not be ignored considering the absence at the time of a treatment or vaccine. One HCW explained: ‘I respect what people say, we should not say that it’s not true. Because we do not have a classic treatment [biomedical treatment] we are told to prescribe Chloroquine. For Westerners, we have to give Azithromycin, Paracetamol against fever, headaches and so on […] You accept what people say and then you advise preventive measures until we have a vaccine’ (IDI 20).

In addition, one participant mentioned prayer as means of insurance against this disease: ‘we must believe in God and pray that the disease does not attack us’ (IDI 7).

#### Preparedness in Faranah and at the Faranah Regional Hospital

All the participants in this study attested that there was a lifestyle change in Faranah following the declaration of the country’s first COVID-19 case. This change was centered on the enforcement of the preventive measures, the closing of worship and leisure places, and the prohibition of social gatherings. For one HCW, these restrictive measures were communicated ‘through the media, radio, and even awareness-raising campaigns. Groups of people went from neighborhood to neighborhood to sensitize people on how to wear masks, even distributing handwashing kits and masks’ (IDI 1).

Participants stated that they observed some changes at the Faranah Regional Hospital. They reported that a triage system and a hand washing station were put in place at the hospital’s main entrance and wards. Some days later, they hired security guards to enforce preventive measures, such as including mask wearing and hand hygiene as a condition for entering the hospital grounds.

Another change was the decrease in the number of HCWs in each service (in particular the ones without contract with the Guinean State) as well as the cessation of the trainees’ work to avoid overcrowding. Participants also described a decrease in the patients’ attendance rate. People sought healthcare via pharmacies or through non-biomedical treatments resulting in a perceived increase in the mortality rate in the hospital as patients were admitted late with complications. Participants emphasized that the slowdown of activities at the hospital had an economic impact on them since HCWs receive a share of the hospital’s revenues from patients’ admissions and treatments at the end of each month.

Concerning triage and treatment, HCWs consistently articulated that there was confusion about suspect case definitions. Some claimed not receiving training about this new virus and treatment, while only a few mentioned both the case definition and treatment. Since the symptomatology resembles other diseases endemic to the region, one HCW reported: ‘in our country, often, the diseases have common signs: it is fever, headache. Now, sore throat and cough are more specific for COVID-19. That is, in addition to all the malaria signs that we usually know, if there is cough and sore throat, and if the patient has lived in an area or has been in contact with a COVID-19 case, it is a suspect case’ (IDI 20).

#### Perception of the preparedness in and outside of the hospital

Most participants reported to rely on the preventive measures enforced at the hospital and the city. However, they did not feel entirely safe because of patients’ lack of compliance. Many participants described a marked difference between the city and the hospital:

> ‘the difference is that in the hospital, we wear gloves and masks, we wash our hands, we wear the complete PPE for example in the lab; but in the city, we don’t wear the gloves, we don’t wear the gown, we only wear masks and wash our hands’ (IDI 1).

This creates a certain distrust between HCWs, patients and community members since the former only feel protected and able to comply with the preventive measures inside the hospital’s facilities. As another HCW argued: ‘in the hospital, all the measures are there, the distances are respected, but in the city, you can’t, for example we women who attend the markets. When we go to the market with the masks, some people look at us badly and you can’t keep the distance’ (IDI 10).

Other participants reported that the city population started neglecting the preventive measures as there were no recorded new cases in the city. Additionally, it is interpreted as conspicuous the perceived failure of the Guinean government to enforce in due time the containment measures and to utilize the most appropriate communication channels. One participant explained:

> ‘In the community, practically, we must refer to the community leaders [for risk communication and community engagement]: people who are loved, who are listened to, who have the confidence of the population, are the ones who should give them this knowledge. […]. Educational leaders [information multipliers] used to come from Conakry, they have the tenure, they go to the prefectures, that is *mamaya* [nonsense] […] We have to go to the communities and choose the key persons there and train them and they will explain to their relatives […] When we do that, we will have good results. Not everyone watches TV. In the villages, people don’t have a TV. The radio, not everyone listens to’ (IDI 5).

Participants who had been in contact with cases in the hospital reported that a kind of mistrust developed between them and their colleagues in other departments who avoided them out of fear of getting infected. One staff member described this as peers’ stigmatization.

### Experiencing COVID-19

We define an experience as a practical contact with and observations of facts and events (49). This section describes cultural and social reactions and the parallels established with the 2014 – 2016 EVD outbreak.

#### Pandemic response in Faranah

HCWs appraised the declaration of the first positive case in the city of Faranah as the most significant moment. One HCW described: ‘the moment that struck me the most was when a patient left another country for Conakry; there, his test was done but he fled to come to Faranah. This panicked the entire population’ (IDI 9). For the participants, there was a community reaction to counteract the virus spread. As one participant put it: ‘In Faranah […] all households had hand washing stations and everyone wore masks. In addition to that, there was the military who monitored the community itself. Everyone was involved so that either you wear your mask because you want to avoid the disease or you wear it because you want to avoid getting fined’ (IDI 4).

Other participants reported a mistrust between the citizens who avoided getting closer to each other for fear of getting infected. One HCWs explained: ‘Yes, there were changes, there was a crisis of confidence between people. Even in vehicles, people did not approach each other and mosques were closed. It altered a lot the social relationships, because people were afraid […] people were running away from each other, the contacts decreased a lot’ (IDI 20).

#### Community reactions

Participants reported disease denial as the first community reaction, following the confirmation of a positive case in the city. Participants explained that there was subsequently an attempt to demonstrate against HCWs accusing them of falsely diagnosing their relatives as positive cases. One of the participants explained:

> ‘When the first case was declared, the population had a negative reaction. First, they hid the person, they didn’t give clear information and they said that because he came from the whites, they [the authorities] thought he had money, that’s why they are looking for him, because he came from Spain. And even with the follow-up team, there were stones thrown at them in the neighborhood’ (IDI 5).

Participants explained that the involvement of the town mayor was necessary. He joined the health authorities to sensitize the population on the local rural radio to decrease tensions. Other participants argued that the population felt disappointed because HCWs could not find treatments to this new infection.

HCWs explained that these tensions led to a crisis of confidence between the HCWs and the population as the latter believed the former to be responsible for the virus spread. This resulted in mistrust, fear and the avoidance of the hospital. One participant pointed out that some people were afraid of the thermo-flash thermometer as it was perceived as a positive cases detector, while another testified that the CT-EpI facilities prevented other people from going to the hospital. They feared the infection would spread from the patients in this service.

#### The shadow of EVD

One prominent theme was the 2014 – 2016 EVD epidemic. Participants established many parallels to describe the COVID-19 outbreak. The experience acquired during EVD is a reassuring factor in counteracting this new pandemic. Nonetheless, it gained a disappointing taint since all the institutional structure and preparedness measures created in the aftermath could not prevent the COVID-19 infection from spreading out of Conakry.

Participants reported that the similarities between the EVD epidemic and the COVID-19 pandemic were clinical symptoms such as fever and physical asthenia, and preventive measures such as hand hygiene. One of the participants also emphasized the infectious nature of both diseases, while another mentioned the mistrust of the population towards the hospital during both outbreaks. The participants relayed several differences between the two diseases. They mentioned new preventive measures introduced during the COVID-19 pandemic, mainly, mask wearing. In addition, the participants alluded to the modes of contamination and the significance of these diseases by referring to the mortality rates.

Despite the high contagiousness of COVID-19 (referring to its rapid spread around the world), most participants agreed that COVID-19 remains less severe than EVD because of its seemingly low mortality rate in the country. However, two other participants reported that it had killed more people worldwide than EVD. In addition, some participants alluded to community reactions when comparing the two diseases. For example, they asserted that there were more violent community reactions during the EVD outbreak because ‘COVID-19 started at the high level of the worldwide authorities, that is to say: “among the whites” before it happened to us here. It came to our country and it started in the ministries, in the directorates before it came to the neighborhoods until today when we talk about community cases’ (IDI 13). Another HCW added: ‘in my opinion, with Ebola it was the fact that people were going to spray the houses. Especially the spraying of the cities, that’s what caused enough problems during Ebola. People thought that the spraying was what made the disease worse’ (IDI 1).

## Discussion

Our study explores the therapeutic landscape of the Faranah Regional Hospital through its HCWs’ beliefs, practices and experiences vis-à-vis the COVID-19 pandemic. The aim is to expand the analytical scope in order to highlight how this pandemic, the preparedness and response plans and resulting governance consequences are experienced, understood and constructed during the hospital’s daily routines. The hospital is thus shaped into a meaningful social space by the experiences of the people who use it the most. This social significance suggests that the HCWs’ understandings of a situation are a good indicator of the societal values and views about this pandemic since their information can measure impacts on access to healthcare services and implementation of preparedness and response measures (50). This is essential to create a people-centered approach to prevention, preparedness and response and to guide by identity leadership.

### 1. Entanglement with the community

A focus on some features of this therapeutic landscape has revealed that HCWs’ knowledge is not constrained to official and/or scientific information as it includes rumors and non-biomedical treatments used by the population. The fact that HCWs rely on different sources to make sense of the pandemic demonstrates that the medical practice is dependent of economic status and social capital (51). HCW’s entanglement with the communities with whom they work and live highlights how porous the barrier between the biomedical world and other social worlds can be. This becomes clear not only by observing the HCWs’ knowledge of non-biomedical treatments, but also because they find explanations for the pandemic in religious worldviews. Similarly, the virus spread and response measures deployment are interpreted in line with the country’s political situation. This influences how the reliability of information sources is perceived (international sources versus national sources).

Research has identified a porosity between social and biomedical worlds in similar settings (37, 52, 53). In these studies, hospitals and the social interactions that occur within are not differentiated by biomedical and non-biomedical practices. By contrast, these practices are understood as co-constituent from each other: they occur across multiple spaces (internal and external from the hospital) and overlap the epistemologies of the different actors. That is why the actors’ relationships change over time and shift their configuration as part of syncretic practices. Our findings confirm this multiplicity and thus strategies to prepare and respond to pandemics need to incorporate it since HCWs appropriate international, national and local health regulations while embodying community perceptions.

### 2. Perceived community differentiation

Another aspect that the therapeutic landscape brings upfront is that HCWs perceive themselves as different from the communities they serve. This differentiation lies in the perceived high value of the knowledge they, as hospital staff, consider to have. This leads to unease and distrust vis-à-vis patients and community members, in particular when the pandemic progression in the country and the city is accounted for. HCWs describe the community as believing rumors and conspiracy narratives to explain the pandemic origins and the use of non-biomedical treatments to counteract the disease. Some even argue that the community’s illiteracy level is the underlying reason for these beliefs and nonbiomedical practices. When it comes to vulnerability perception, HCWs interviewed in this study also draw a clear difference between their routines at the hospital and the city and/or the community. They argue that in the former all protective measures are enforced and complied with, while in the latter there is a relaxation which prevents the HCWs from complying and puts them in danger.

Psychological and sociological literature has highlighted that pre-existing group identities, norms, values, and worldviews affect people’s (non-)compliance with public health advice (27). This literature has also shown that in-group and out-group memberships have affected vulnerability perceptions during the COVID-19 pandemic (27, 54). Research on HCWs’ identity in African settings has stressed that HCWs have to constantly negotiate and re-negotiate their identity with other social roles that are seemingly conflicted but potentially complementary (26). In our study, variations between different HCWs identities (or professions and/or services) did not emerge as a significant point of tension except with colleagues who cared for the few COVID-19 positive patients at the hospital. Nonetheless, the differentiation is particularly salient between being a HCW (in-group identity) and other members of the community (out-group). Generally, a strong lack of trust towards the healthcare system and social resistance to response teams has been identified during epidemics, but not from HCWs to communities (25, 55).

### 3. Experiencing the pandemic is not the same as responding to it

Our results show a strong community embeddedness of the HCWs. In addition, we extrapolate a strong in-group and out-group distinction regarding the community. This pinpoints the difference between what is believed and experienced and what is practiced. Consequently, the elevated risk posed by in-group members should be recognized in preparedness and response plans since most people could find it easier to grasp the risk posed by a stranger than by a colleague (56). This differentiation between perception and practice seems also to influence mental health of HCWs (particularly risk perception and the stress associated when the risk is perceived as high) and healthcare provision during outbreaks (30). Our findings underline as well how HCWs mobilize, negotiate and translate different practices, discourses and identities to make sense of their lifeworld (41). Even if it seems contradictory, accounting for the complexity within the beliefs, the experiences and the practices of HCWs is of great importance for the understanding of how preparedness and response are locally acted upon under particular infrastructural, technical and social pressures (41, 57).

### 4. People-centered approaches and other preparedness and response measures should become operational

The 2021 WHO Strategic Preparedness and Response Plan for the WHO African Region recommended the management of the infodemic (an overabundance of information, including misinformation) by ensuring access to evidence-based information at the right time from trusted sources. The plans emphasize as well safety and security of frontline HCWs in the national preparedness and response plan for COVID-19 (9). These guidelines state that African countries will need to move towards people-centered and community-led approaches to the response, which will increase trust and social cohesion. Moreover, lessons learned from the COVID-19 pandemic accentuate community involvement hand-in-hand with measures to resolve systemic issues and the management of social resistance and rumors without recurring to violent enforcement (5). They also accentuate the need to recruit local staff in the response, including local people (e.g. grass-root and influential community leaders) to build the response structures.

Our findings confirm that poor access to quality health services persists (e.g. lack of clarity about laboratory results and low reliability of antigen rapid tests). Our results demonstrate as well that tensions continue around rumors management and communicating officially updated information about the pandemic to HCWs and other relevant actors of the healthcare system. We underline therefore the importance of managing infodemics as an essential tool in prevention, preparedness and response of outbreaks. Infodemics management should encompass social media and also traditional media like the radio which is an essential information source in African societies. In Guinea, research shows that radio conveys messages which translate/appropriate global information while mobilizing epidemic experiences embedded in the political and social history of the country (58).

In addition, the community reactions encountered by the HCWs and the description of the preparedness measures enforcement in the city and at the hospital suggest a low active involvement of local people (especially local leaders but also community HCWs) for community engagement and building trust bridges between the healthcare system and the community. It should be noted that this lack of trust and miscommunication can have historical roots, which revive with every epidemic and at each peak of social tension making local leadership to be contingent on each place and situation (59). More efforts are therefore required to include people-centered approaches that account for this contingency. Similarly, more research is needed to understand why such approaches did not become operational, even though they have been highlighted as fundamental during the management of previous epidemics and of other global health emergencies (60–63).

## Strengths and limitations

The study addresses a relevant research gap in health emergency preparedness, prevention and control. It focuses on building bridges between people-centered preparedness and response plans and the way they are acted upon by one of the most relevant stakeholders - the HCWs. Our findings should, nonetheless, be analyzed considering some limitations. We formulated the themes discussed in this paper in the context of a broader study on pandemic preparedness response via a HCWs’ Knowledge, Attitudes and Practices research. As such, our study was not designed to assess how HCWs’ beliefs, practices and experiences change over time as the pandemic evolves. Furthermore, other groups that intrinsically belong to the hospital’s therapeutic landscape, such as patients, maintenance staff, caregivers and families were not studied. Similarly, this study did not investigate gender, age, professional experience and ethnicity as variables, and their connection with mistrust and risk perception (34, 43, 64).

## Conclusion

Our study aims to further deepen the understanding of the COVID-19 pandemic in Guinea, starting with healthcare facilities but seeking to explore aspects which transcend the hospital itself and speak about broader processes encompassing social and political phenomena as well as national and international sanitary regulations. Literature about outbreak preparedness emphasizes epidemiological surveillance via the strengthening of community-led initiatives. We argue that, within these plans, in order to effectively incorporate people-centered approaches, more structural efforts are needed to incorporate the social landscapes of the disease, including 1) places (like hospitals) where different social tensions related to the outbreak are particularly salient and 2) people (HCWs) who are predominantly vulnerable and with whom engagement should start. For a more comprehensive outbreak response, we recommend to account for shifting and simultaneous identities from HCWs as well as overlapping realities about how a pandemic is understood, prioritized, perceived and experienced in order to reduce tensions around rumors management and communication of officially updated information.

## Data Availability

The data sets generated and/or analyzed during the current study are not publicly available because data has been collected from a small group of participants, and even data that are not directly identifying in combination becomes identifying (e.g. sex, profession, department ward, work description). Data is nonetheless available from the corresponding author on reasonable request.

## Acknowledgements

We would like to thank all study participants at the HRF for openly answering our questions and explaining to us their routines and impressions. We would also extend our most sincere appreciation to the Global Health Protection Program (GHPP) of the German Ministry of Health (Bundesministerium für Gesundheit) for funding the project (Grant code: ZMVI1-2519GHP710). Special thanks to Dr. Alpha Oumar Karim Diallo for his collaboration, openness and commitment, and to Dr. Matthias Borchert, Dr. Sophie Müller and Dr. Seth Kofi Abrokwa1for critically reviewing an earlier version of the manuscript.

## Abbreviations

ANSS: Agence Nationale de Sécurité Sanitaire
CT-EPI: Center for the Treatment of Epidemics
EVD: Ebola Virus Disease
FRH: Faranah Regional Hospital
HCW: Healthcare workers
IDI: Semi-structured interview

## References

1. Martineau F, Wilkinson A, Parker M. Epistemologies of Ebola: Reflections on the Experience of the Ebola Response Anthropology Platform. Anthropological quarterly. 2017;90(2):475–94.

2. Bolten C, Shepler S. Producing Ebola: Creating Knowledge In and About an Epidemic. Anthropological quarterly. 2017;90(2):349–68.

3. Venables E, Pellecchia U. Introduction: Engaging Anthropology in an Ebola Outbreak: Case Studies from West Africa. Anthropology in action (London, England : 1994). 2017;24(2):1.

4. Sharon Abramowitz JB. Responding to Ebola: creating an agile anthropology network. 2016.

5. Grant C, Achyut P, Akello G, Alam E, Ayegboyin M, Baluku M, et al. People’s Agenda for Pandemic Preparedness. Institute of Development Studies. 2023.

6. Bedford J, Farrar J, Ihekweazu C, Kang G, Koopmans M, Nkengasong J. A new twenty-first century science for effective epidemic response. Nature (London). 2019;575(7781):130–6.

7. Giles-Vernick T, Kutalek R, Napier D, Kaawa-Mafigiri D, Dückers M, Paget J, et al. A new social sciences network for infectious threats. The Lancet Infectious Diseases. 2019;19(5):461–3.

8. Carter SE, Ahuka-Mundeke S, Pfaffmann Zambruni J, Navarro Colorado C, van Kleef E, Lissouba P, et al. How to improve outbreak response: a case study of integrated outbreak analytics from Ebola in Eastern Democratic Republic of the Congo. BMJ Global Health. 2021;6(8):e006736.

9. WHO-Afro. The Coronavirus disease 2019 (Covid-19): Strategic Preparedness and Response Plan for the WHO African Region. 2021.

10. Cristea F, Weishaar H, Geurts B, Delamou A, Tan MMJ, Legido-Quigley H, et al. A comparative analysis of experienced uncertainties in relation to risk communication during COVID19: a four-country study. Globalization and health. 2022;18(1):66.

11. Weishaar H, Geurts B, Mari-Saez A, Delamou A, Jegede A, Legido-Quigley H, et al. Risk communication during COVID-19: a comparative analysis of four countries. European Journal of Public Health. 2021;31(Supplement_3).

12. MwFRHP. Coronavirus in Africa: New variants are causing growing concern. BBC news. 2021.

13. Sankoh O, Dickson KE, Faniran S, LahFRHJI, Forna F, Liyosi E, et al. Births and deaths must be registered in Africa. The Lancet Global Health. 2020;8(1):e33–e4.

14. Okonji EF, Okonji OC, Mukumbang FC, Van Wyk B. Understanding varying COVID-19 mortality rates reported in Africa compared to Europe, Americas and Asia. Tropical medicine & international health. 2021;26(7):716–9.

15. MacGregor H, Leach M, Akello G, Sao Babawo L, Baluku M, Desclaux A, et al. Negotiating Intersecting Precarities: COVID-19, Pandemic Preparedness and Response in Africa. Medical anthropology. 2022;41(1):19–33.

16. Kolié D, Keita FN, Delamou A, Dossou J-P, Van Damme W, Agyepong IA. Learning from the COVID-19 pandemic for future epidemics and pandemics preparedness and response in Guinea: Findings from a scoping review. Frontiers in Public Health. 2022;10.

17. Bonnet E, Bodson O, Le Marcis F, Faye A, Sambieni NE, Fournet F, et al. The COVID-19 pandemic in francophone West Africa: from the first cases to responses in seven countries. BMC public health. 2021;21(1):1490.

18. World Health O. Coronavirus disease 2019 (COVID-19): situation report, 62. Geneva: World Health Organization; 2020 2020-03-22.

19. MoH-ANSS. Plan National de Préparation et de Riposte contre l’Épidémie de Coronavirus (CoVid-19). 2020.

20. MoH-ANSS. Guinée - Covid 19 - Rapport de situation 13 (1 juillet 2020). 2020.

21. Université Gamal Abdel Nasser de Conakry C-P, ENABEL. De l’épidémie d’Ébola à la pandémie de Corona virus: une Analyse situationnelle en Guinée. 2020.

22. Delamou A, Sow A, Fofana TO, Sidibé S, Kourouma K, Sandouno M, et al. A rapid assessment of health system preparedness and response to the COVID-19 pandemic in Guinea. J Public Health Afr. 2022;13(2):1475.

23. Carrión Martín AI, Derrough T, Honomou P, Kolie N, Diallo B, Koné M, et al. Social and cultural factors behind community resistance during an Ebola outbreak in a village of the Guinean Forest region, February 2015: a field experience. International health. 2016;8(3):227–9.

24. Thiam S, Delamou A, Camara S, Carter J, Lama EK, Ndiaye B, et al. Challenges in controlling the Ebola outbreak in two prefectures in Guinea: why did communities continue to resist? The Pan African medical journal. 2015;22 Suppl 1(Suppl 1):22.

25. Fairhead J. Understanding Social Resistance to the Ebola Response in the Forest Region of the Republic of Guinea: An Anthropological Perspective. African studies review. 2016;59(3):7–31.

26. Mlotshwa L, Harris B, Schneider H, Moshabela M. Exploring the perceptions and experiences of community health workers using role identity theory. Glob Health Action. 2015;8:28045-.

27. Smith LGE, Gibson S. Social psychological theory and research on the novel coronavirus disease (COVID-19) pandemic: Introduction to the rapid response special section. British Journal of Social Psychology. 2020;59(3):571–83.

28. Aborode AT, Tsagkaris C, Jain S, Ahmad S, Essar MY, Fajemisin EA, et al. Ebola Outbreak amid COVID-19 in the Republic of Guinea: Priorities for Achieving Control. The American journal of tropical medicine and hygiene. 2021;104(6):1966–9.

29. Attas F, Curtis, Marie Ivonne, Koniomo Niouma Laurent, Le Marcis, Frédéric. Covid-19 en Guinée. Impact de l’Hétérogénéité des Soins sur la Perception de la Prise en Charge. Guinée: AFD, IRD, CERFIG; 2021. Contract No.: 8.

30. Afulani PA, Gyamerah AO, Nutor JJ, Laar A, Aborigo RA, Malechi H, et al. Inadequate preparedness for response to COVID-19 is associated with stress and burnout among healthcare workers in Ghana. PLoS One. 2021;16(4):e0250294.

31. Chersich MF, Gray G, Fairlie L, Eichbaum Q, Mayhew S, Allwood B, et al. COVID-19 in Africa: care and protection for frontline healthcare workers. Globalization and Health. 2020;16(1):46.

32. Chemali S, Mari-Sáez A, El Bcheraoui C, Weishaar H. Health care workers’ experiences during the COVID-19 pandemic: a scoping review. Human Resources for Health. 2022;20(1):27.

33. Doumbouya ML. Pauvreté et accessibilité aux services de santé: le cas de la Guinée. Économie et solidarités. 2007;38(2):137–53.

34. Jaffré Y. Une médecine inhospitalière : les difficiles relations entre soignants et soignés dans cinq capitales d’Afrique de l’Ouest / sous la dir. de Y. Jaffré. Paris [u.a: Karthala; 2003.

35. d’Alessandro E. Prévenir le risque infectieux à l’hôpital ?: Réflexions anthropologiques autour des pratiques d’hygiène hospitalière dans un service de médecine au Niger. Anthropologie & santé. 2012(4).

36. Olivier de Sardan J-P, Diarra A, Moha M. Travelling models and the challenge of pragmatic contexts and practical norms: the case of maternal health. Health research policy and systems. 2017;15(Suppl 1):60-.

37. Bonnet D, Jaffré Y, editors. Les maladies de passage : transmissions, préventions et hygiènes en Afrique de l’Ouest. Paris: Karthala; 2003.

38. Zaman S. Poverty and violence, frustration and inventiveness: hospital ward life in Bangladesh. Soc Sci Med. 2004;59(10):2025–36.

39. van der Geest S, Sarkodie S. The fake patient: A research experiment in a Ghanaian hospital. Social Science & Medicine. 1998;47(9):1373–81.

40. Ashworth HC, Dada S, Buggy C, Lees S. The Importance of Developing Rigorous Social Science Methods for Community Engagement and Behavior Change During Outbreak Response. Disaster Med Public Health Prep. 2020:1–6.

41. Attas F. Epidemics in Guinea: from infectious politics to viral ontologies. À propos. 2022:109.

42. Madge C. Therapeutic landscapes of the Jola, The Gambia, West Africa. Health & Place. 1998;4(4):293–311.

43. Leach MA, Fairhead JR, Millimouno D, Diallo AA. New therapeutic landscapes in Africa: Parental categories and practices in seeking infant health in the Republic of Guinea. Social Science & Medicine. 2008;66(10):2157–67.

44. Winchester M, McGrath J. Therapeutic landscapes: Anthropological perspectives on health and place. Medicine Anthropology Theory | An open-access journal in the anthropology of health, illness, and medicine. 2017;4(1).

45. Douno M, Rocha C, Borchert M, Nabe I, Müller SA. Qualitative assessment of hand hygiene knowledge, attitudes and practices among healthcare workers prior to the implementation of the WHO Hand Hygiene Improvement Strategy at Faranah Regional Hospital, Guinea. PLOS Glob Public Health. 2023;3(2):e0001581.

46. Doughty K, Hu H, Smit J. Therapeutic landscapes during the COVID-19 pandemic: increased and intensified interactions with nature. Social & cultural geography. 2022;ahead-of-print(ahead-of-print):1–19.

47. Gruénais M-E, Pourtier R, editors. La santé en Afrique : anciens et nouveaux défis 2000.

48. Vivanco LA. A Dictionary of Cultural Anthropology. Online: Oxford University Press; 2018.

49. Oxford. Oxford English Dictionary: Oxford University Press; 2021.

50. Groups AfOC-RRSSW. Guidance for Health Care Worker (HCW) Surveys in Humanitarian Contexts in LMICs. Unicef, WHO, Goarn and others; 2021.

51. Fairhead J, Leach M, Small M. Where techno-science meets poverty: medical research and the economy of blood in The Gambia, West Africa. Social science & medicine (1982). 2006;63(4):1109–20.

52. Street A, Coleman S. Introduction Real and Imagined Spaces. Space and Culture. 2012;15:4–17.

53. Chabrol FaKJ. The Hospital Multiple: Introduction. Somatosphere. Online 2020.

54. Philipps J, Sagnane S. A Disease of the Rich and a Disease of the Poor. 2023. p. 128–42.

55. LeMarcis F, Enria L, Abramowitz S, Saez A-M, Faye SLB. Three Acts of Resistance during the 2014–16 West Africa Ebola Epidemic. Journal of Humanitarian Affairs. 2019;1(2):23–31.

56. Cruwys T, Stevens M, Greenaway KH. A social identity perspective on COVID-19: Health risk is affected by shared group membership. British Journal of Social Psychology. 2020;59(3):584–93.

57. Wolf M, Hall K. Cyborg Preparedness: Incorporating Knowing and Caring Bodies into Emergency Infrastructures. Medical anthropology. 2018;37(6):486–98.

58. Attas F, Keïta-Diop M, Curtis M, Le Marcis F. Discours radiophoniques, cartographies épidémiques et représentations locales de la COVID-19 en GuinéeRadio discourses, epidemic maps and local representations of COVID-19 in Guinea. L’Espace Politique. 2022;44.

59. Fribault M. Ebola en Guinée : violences historiques et régimes de doute. Anthropologie & santé. 2015.

60. Abramowitz SA, McLean KE, McKune SL, Bardosh KL, Fallah M, Monger J, et al. Community-centered responses to Ebola in urban Liberia: the view from below. PLoS neglected tropical diseases. 2015;9(4):e0003706-e.

61. Biehl J, Petryna A. When people come first: critical studies in global health. STU - Student edition ed. Princeton: Princeton University Press; 2013.

62. Alain Epelboin AO-KaPF. Contribution de l’anthropologie médicale à la lutte contre les épidémies de fièvres hémorragiques à virus Ebola et Marburg. 2014.

63. Farmer P. Social inequalities and emerging infectious diseases. Emerg Infect Dis. 1996;2(4):259–69.

64. Jaffré Y, Lange IL. Being a midwife in West Africa: Between sensory experiences, moral standards, socio-technical violence and affective constraints. Social science & medicine (1982). 2021;276:113842.

